# What are the ethnic inequities in care outcomes related to haematological malignancies, treated with transplant/cellular-therapies, in the UK?

**DOI:** 10.1101/2024.11.18.24317089

**Authors:** Samuel Cusworth, Zareen Deplano, Alastair Denniston, David Burns, Krishnarajah Nirantharakumar, Nicola J Adderley, Joht Singh Chandan

## Abstract

**Introduction:** Haematological cancers are common in the UK, with a variety of morphologies. Stem cell transplants and chimeric antigen receptor (CAR) T-cell therapies provide significant options for hard to treat haematological cancers, although with difficult to predict outcomes. Research into the determinates of treatment efficacy, and access to treatments, is key to ensuring equal benefit across patients, and patient safety. With this, there are concerns about the small representation of minority groups in related research. We aim to report on the current knowledge, to guide future research.

**Methods and analysis:** A variety of databases will be searched, for literature on UK minority ethnic populations receiving haematopoietic stem cell transplant or CAR T-cell therapy. Many outcomes will be analysed, covering the patient care pathway for those of the target population, although with a focus on follow-up after therapy. Plans have been made to conduct narrative synthesis, with meta-analysis where applicable.

**Ethics and Dissemination:** Outputs will be published in an appropriate journal, and discussed with the wider NIHR Blood and Transplant Research Unit in Precision Transplant and Cellular Therapeutics (BTRU) group. Discussions will also be undertaken with the BTRU patient partners group.

**Strengths and Limitations of this Study:**

- This systematic review has a detailed search criteria with a variety of search tools, enabling high sensitivity in obtaining evidence.
- To enable this work to be completed, reviewers will be working independently on separate reference lists, during screening, rather than voting on the same references and resolving conflicts. To alleviate issues, reviewers will be first required to screen a defined number of papers together, using a more classical approach to confirm agreement, before working independently.
- Due to difficulties in pooling together international ethnic groups, only studies in the UK have been included, to reduce complexity. There are some ethnic groups which are more valid to pool together internationally than others, mainly in respect to genetic factors (as opposed to social), which this protocol does not consider.

## Introduction

### Inequities in haematological cancer outcomes

Haematological cancers are the fifth most common cancer in the UK as of Aug-2022 (1). They cover a variety of different cancers, diagnosed by the cells effected, the location and morphology, with the broad classifications of leukaemias (leukocytes), myelomas (myeloid cells) and lymphomas (lymphocytes). Stem cell transplants and CAR T-cell therapies are important in the treatment of relapsing and refractory haematological cancers, providing options in cases difficult to treat. The number of stem cell transplants administered in the UK is increasing; the British Society of Blood and Marrow Transplantation and Cellular Therapy (BSBMTCT) reports an average 5% increase per year of transplants (2006-2019) (2). Despite the positive impact of these treatments, it remains difficult to predict treatment outcomes for haematological cancers, such as cytokine release syndrome, neurotoxicity, and cancer relapse (3). Understanding the factors determining responses to treatment, across applicable blood cancers, is key to improving patient care. To achieve this, trials/studies need to report results that are reflective of the entire population, to prevent disparities in outcomes.

Studies on haematological cancer clinical trials show evidence of under-representation of different ethnic minority populations (4). A study conducted on Asian paediatric patients found evidence of lower survival in lymphoma cases (although this could not be confirmed due to high correlation with area deprivation) (5). This study emphasised the need for better ethnicity and sociodemographic data to measure these inequities in outcomes. A study in black and white patients with multiple myeloma found greater tumour mutational burden for the black ethnic group than was true, due to the lack of representation in public genomic data (6). This makes the test for these mutations potentially cause racial bias across patients. A USA study looking at survival rates of multiple myeloma, highlighted improving survival rates over time, although basing these conclusions mainly on the white population (7). This study found that the white population showed the greatest temporal improvement of survival, compared to other ethnic groups. This demonstrates research potentially masking important health disparities. Furthermore, a study looking to predict risk of poor outcomes across those with diffuse large B-cell lymphoma treated with CAR T, reported that ethnicity/race was not available for the analyses (3). This assumes ethnicity does not effect risk predictions here, masking potential issues if this assumption is not met.

### Policy and role of research in addressing inequalities

It is important for analysis of variations in patient care to be fully representative of the patient population. Inappropriate representation/analysis of patient subpopulations can mask inequalities in care and reduce the ability to make informative decisions to improve on this. Due to data bias towards those of white populations, much clinical research is based around this over-representation. A systematic review on AI prediction methods for breast cancer, found that a majority of analysed datasets under-reported ethnicity, and from those reported, a large bias towards white populations was found (8). It is acknowledged that strategies to reach the white populations are not as effective at reaching other minority ethnic groups (4,9). Therefore, it is important to consider a range of methods when conducting studies or designing strategies to reach these groups, to ensure appropriate coverage of all populations. For example, where a population has shown loss of trust in healthcare organisations, ensuring communication comes from a source as deemed trustworthy by this population, with appropriate terms used to avoid negative connotations, can improve participation (4).

Research into the involvement/inclusion of minority ethnic groups in clinical trials, in the UK, is scarce (4). Current and past UK research has/had limited enforcement of diversity in research, using guidance, rather than making this a mandatory inclusion (as in the US) (10). The Care Act 2014 (11) introduced the Health Research Authority (HRA) (12), amending this into the public authorities listed in the Equality Act 2010 (13). Project-based, England-led, NHS Health and Social Care research requires HRA approval. The HRA ensure research is ethical, in which they provide guidance to research ethics committees to ensure they assess diversity in research projects. They are currently reviewing and looking to update this guidance (12). The HRA are also looking at updating the UK policy framework for health and social care research, to make the importance and expectation of diverse research more explicit. Regarding health and social care in the UK, the Health and Care Act of 2022 (14) and Health and Social Care Act of 2012 (15), made legislations to improve the equality of these services. NHS England, alongside other public authorities, have legal duties to ensure equality is monitored and improved upon. Overall, much guidance is becoming available to improve diversity in health research, with potential for clearer legislation regarding this in the years to come.

### Addressing Issues

Despite the broader evidence in the US compared to the UK, Kirtain et al (2017) still reports the lack of evidence regarding outcomes, across different USA ethnic groups, for haematological cancers, when compared to solid tumours (16). Furthermore, due to differences in the healthcare infrastructure in the UK, compared to the USA, it is important to analyse outcomes relative to the UK. This supports the need for the collection/generation of up-to-date evidence on barriers to the treatment of minority populations in haematological cancer, and their involvement in research. This follows a recommendation from the Anthony Nolan trust (17) - ‘More research is required to gain a better understanding of how factors such as income, education level, social marginalisation, poor quality housing and health literacy affect access to treatment and outcomes; impact stem cell transplant patients’ quality of life and wellbeing; and the unmet needs of these groups.’

### Aims

The primary aim of this review is to describe the breadth of knowledge of inequities experienced by different ethnic groups which occur at all stages in haematological cancer care, for those treated with transplants/cellular-therapies, across ethnic minority groups, in the UK.

### Objectives

1. To describe inequities relating to quality-of-care, co-morbidities and mortality.
2. To describe any known mechanisms contributing to these inequities.
3. Identification of potential areas of systemic bias and weaknesses in study designs which may make them susceptible to not appropriately identifying inequities.

## Methods and Analysis

This protocol was developed in accordance with PRISMA-P guidelines, registered on PROSPERO (ID CRD42024535405) (18,19). The synthesis of quantitative data was planned using guidance from the Cochrane handbook (20). This review will also be undertaken using guidance from the Cochrane handbook, and reported in accordance with PRISMA-P preferred reporting items for systematic reviews and meta-analysis (18,20).

### Eligibility Criteria

Primarily this review will focus on observational studies, which capture information regarding ethnicity.

This systematic review will incorporate the following quantitative study designs (as labeled by Joanna Briggs Institute (JBI) (21)): analytical cross-sectional studies, case-control studies, cohort studies, prevalence studies, systematic reviews and research syntheses, economic evaluations, and text and opinion.

Importantly the review will exclude all studies that do not include a UK population. The meaning of ethnicity and different ethnic groups differs across countries hence to avoid heterogeneity we will focus solely on a UK population. Additionally, the structural differences, and population make-up, between the health care systems and populations (respectively) of different countries and the UK differ. Therefore, only using studies involving UK populations, ensures conclusions will be applicable to the UK population. The inclusion/exclusion criteria are summarised below in table 1. Broadly the PECO for the study is:

**Table 1.** Inclusion exclusion criteria.

| Inclusion | Exclusion |
| --- | --- |
| Population: reports on haematological cancers treated with transplants/cellular-therapies (mentioned above) (irrespective of whether patient has or not). Part of the UK population. | Where outcomes are not presented broken down by ethnic group.<br>Non UK studies |
| Reports on one of the outcomes of interest: Stage of cancer at detection/referral, treatments/interventions received, methods of diagnosis, time intervals (between care pathway stages), screening rates, co-morbidities (risk, burden), survival rates.<br>Must report outcomes for CAR T-cell therapy or haematopoietic stem cell transplant. | Does not mention CAR T-cell therapy or stem cell transplant. |
| Study design one of: analytical cross-sectional studies, case-control studies, cohort studies, prevalence studies, randomised control trials, systematic reviews and research syntheses, text and opinion. | Studies on cellular biology (e.g. looking at gene/cell effect/function, looking at relationship between specific cells and a disease). Do not exclude if reporting on inclusion outcomes, and using mutational markers to disaggregate the data.<br>Qualitative research.<br>Animal studies. |
| Paper available in English. Study published from year 2011 onwards. | Non-English language texts. |

#### Population

Individuals in the UK with haematological cancers treated with transplant/cellular therapies, as described by the British Society of Blood and Marrow Transplantation and Cellular Therapy (BSBMCT) (2,22,23). These haematological cancers are (included where indolent lymphomas have transformed to diffuse large B-cell lymphoma):

- Subset of leukaemias:
  ∘ Acute myeloid,
  ∘ Acute lymphoblastic/lymphocytic,
  ∘ Chronic myeloid,
  ∘ Chronic lymphocytic (or small lymphocytic lymphoma (24)) including prolymphocytic leukemia (24),
  ∘ Plasma cell,
- Hodgkin’s lymphoma/disease:
  ∘ Classical,
  ∘ Nodular lymphocyte predominant B-cell lymphoma (previously called Nodular Lymphocyte Predominant Hodgkin Lymphoma),
- Subset of Non-Hodgkin’s Lymphomas:
  - Diffuse large B-cell (DLBCL),
    ▪ Primary mediastinal large B-cell (separately defined from DLBCL in IDC10 (25,26)),
- Follicular,
- Peripheral T-cell (excluding cutaneous lymphomas),
- Mantle cell,
- Lymphoblastic (or acute lymphoblastic),
- Anaplastic large cell,
- Primary CNS,
- Lymphoplasmacytic (AKA Waldenström macroglobulinemia),
- Small lymphocytic (or chronic lymphocytic leukaemia (24)),
- High-grade/aggressive mature B-cell (previously Burkitt),
- Transformed non-Hodgkin’s Lymphomas:
- Follicular lymphoma,
- Chronic Lymphocytic Leukaemia/Small Lymphocytic Lymphoma (Richter’s syndrome/transformation/lymphoma),
- Nodular lymphocyte predominant B-cell lymphoma (previously called Nodular Lymphocyte Predominant Hodgkin Lymphoma),
- Waldenström macroglobulinemia,
- Marginal zone lymphoma,
- Mucosa associated lymphoid tissue (MALT),
- Myeloma (AKA multiple myeloma),
- Myelodysplastic syndrome (AKA myelodysplasia, myelodysplastic neoplasm),
- Myelofibrosis.

#### Exposure of interest

Minority ethnic groups (defined by UK government as ‘all ethnic groups except for the white British group’ (27)).

#### Comparator

Other ethnic groups.

#### Outcomes of interest

(1) Stage of cancer at detection/referral and mode of presentation, (2) treatments/interventions received, (3) methods of diagnosis, (4) time intervals (between care pathway stages), (5) screening rates, (6) co-morbidities (risk, burden), (7) survival rates.

### Information Sources

With the assistance of an information specialist, the following databases/search-tools will be searched to identify relevant evidence: Cochrane library (28), Epistomonikas (29), Campbell systematic reviews (30), Health evidence (31), PubMed (Medline (32), PubMed central and Bookshelf) (33), OpenGrey (EasyGrey) (34), Ovid Evidence-based Medicine Reviews (EBMR) (searching EMBASE (35) and Medline (32)) (36), Web of Science (37), Scopus (38) and Proquest (39).

### Search Strategy

A comprehensive search strategy (table 2) has been developed with the support of an experienced information specialist.

**Table 2.**
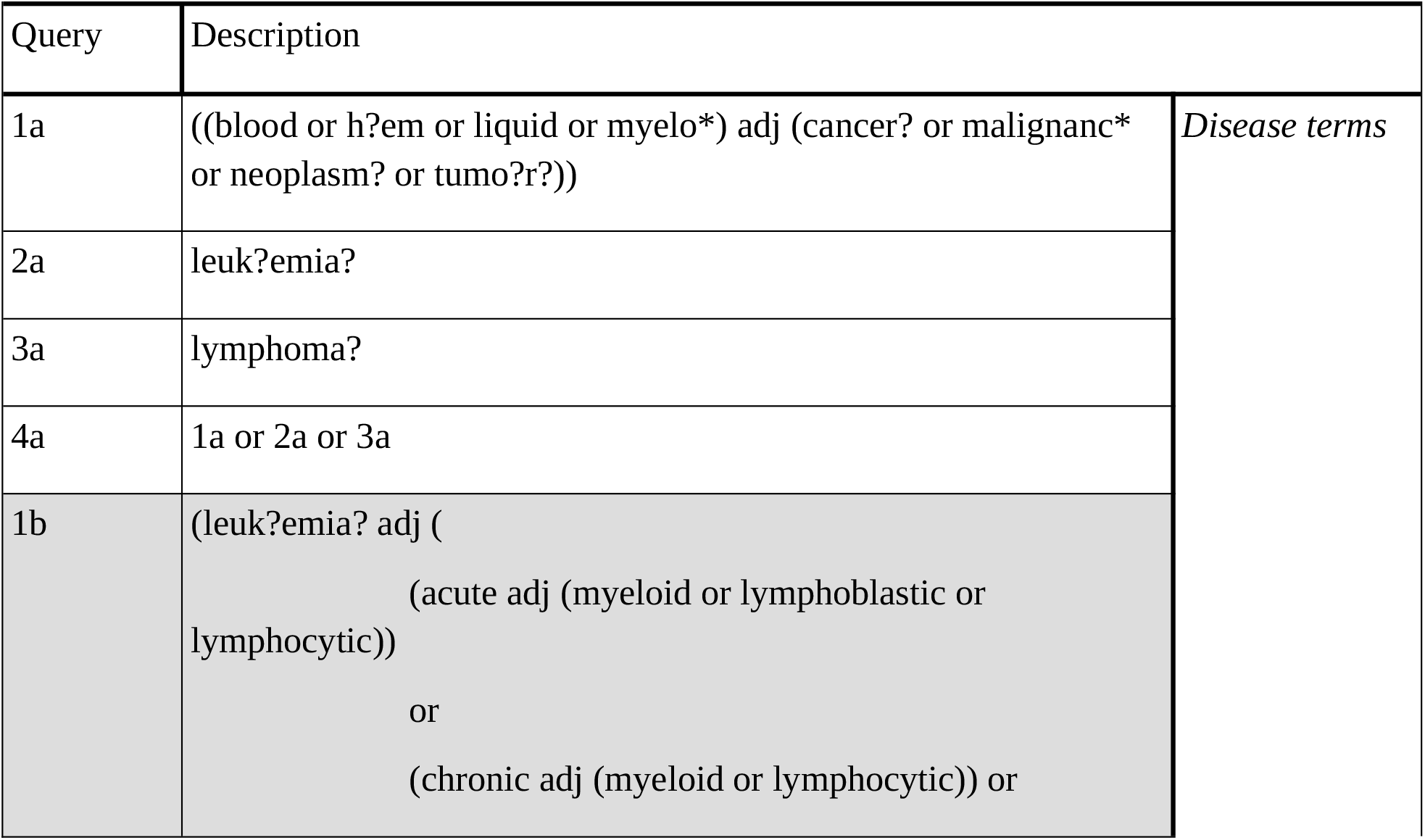
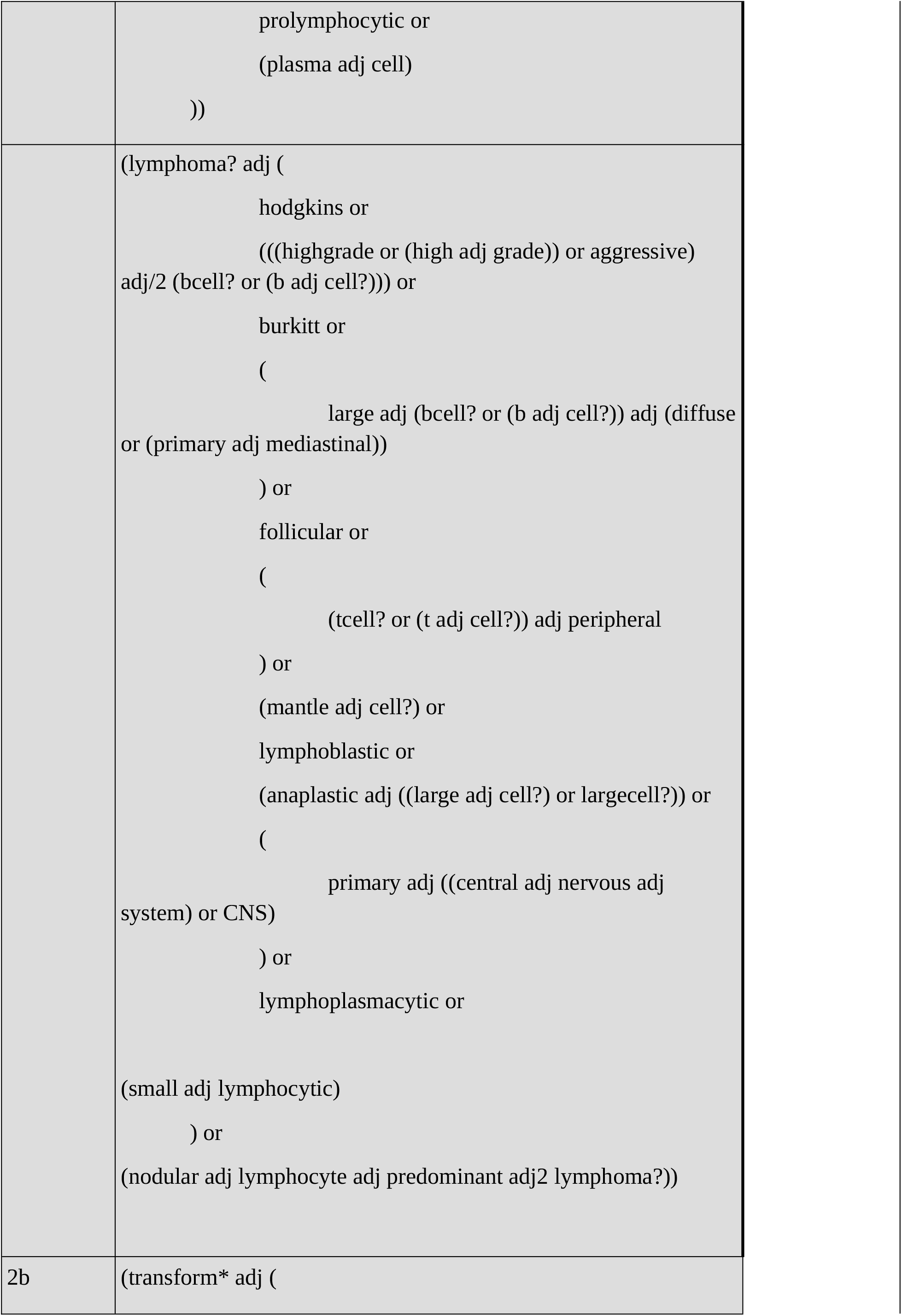
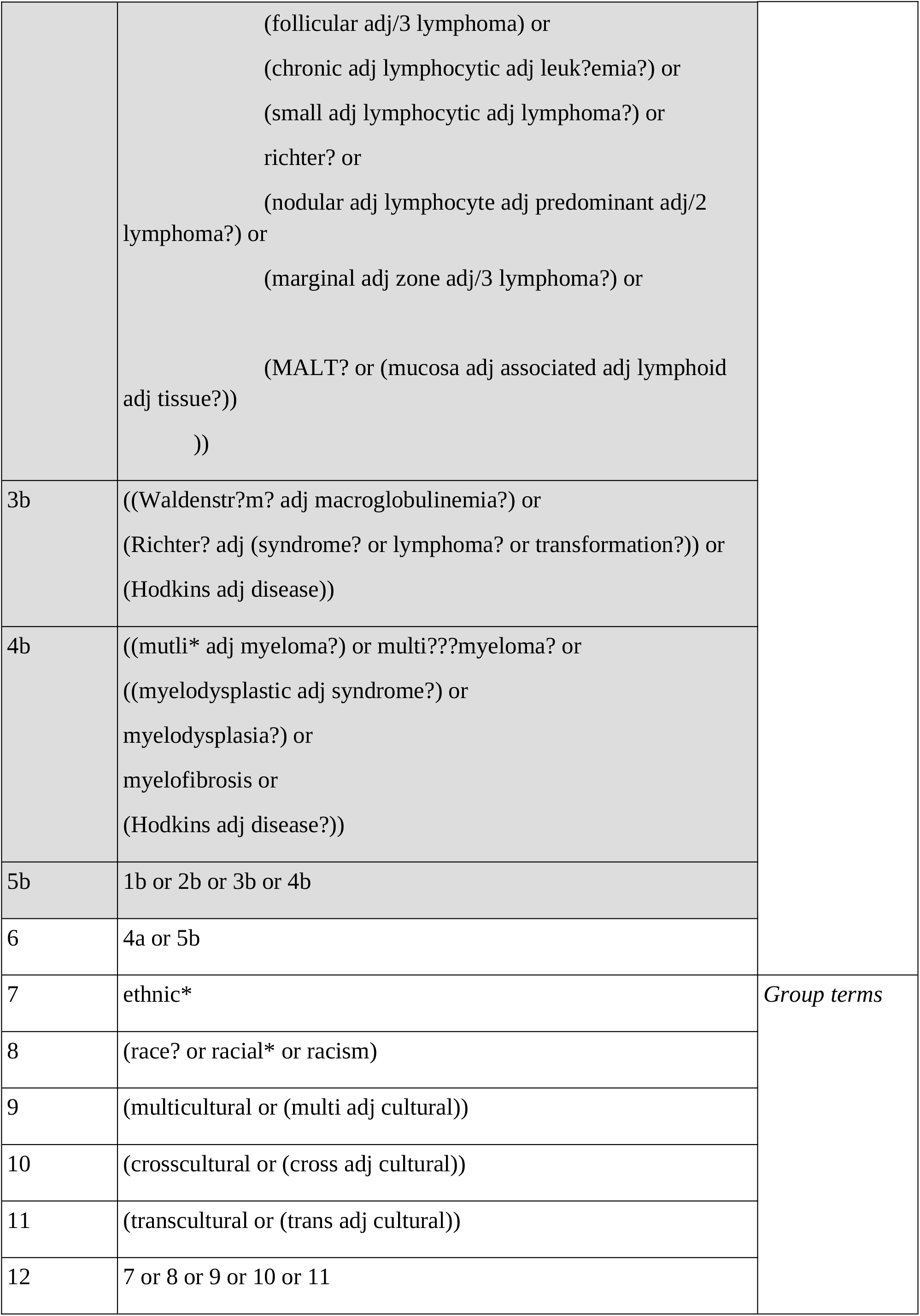
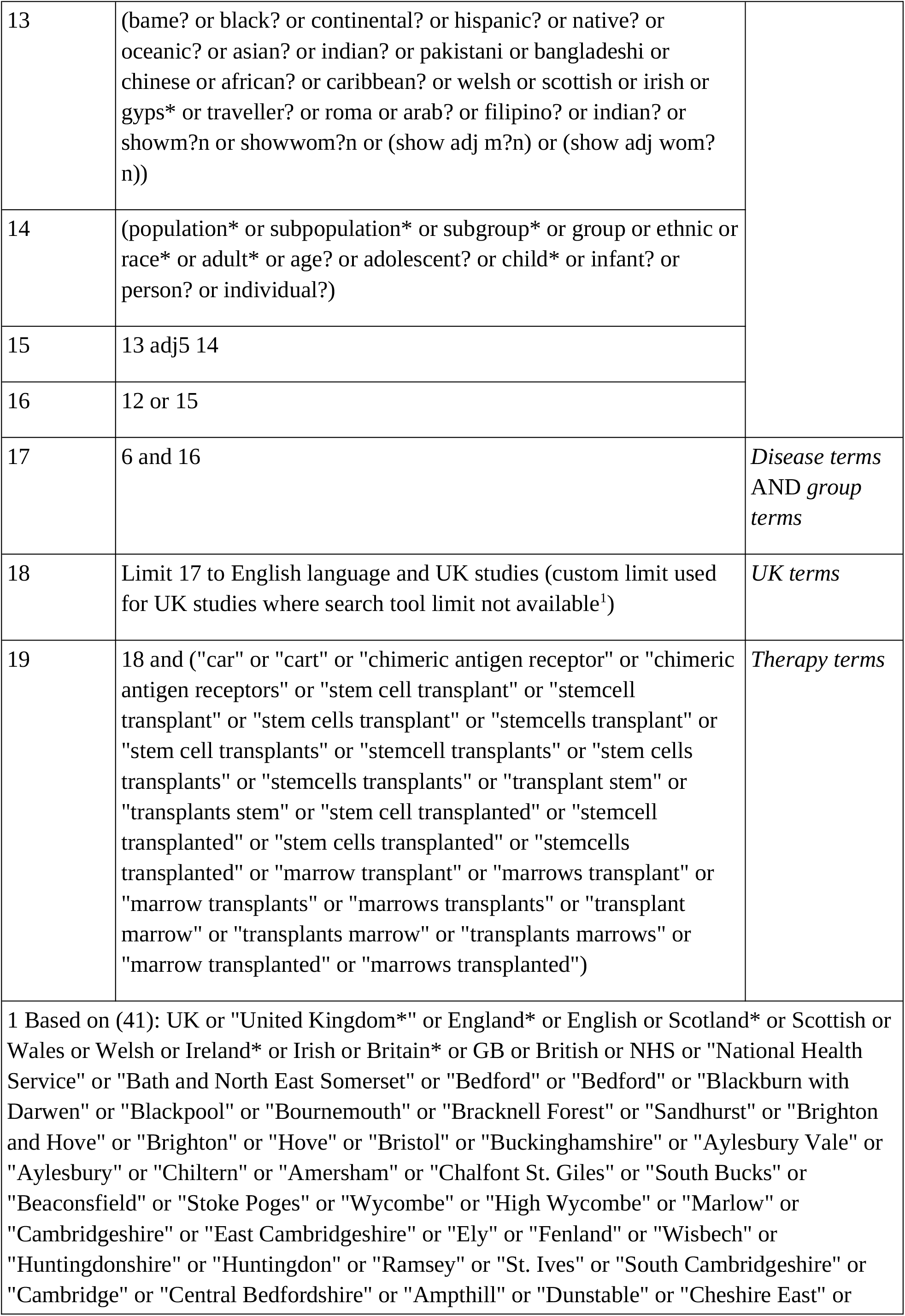
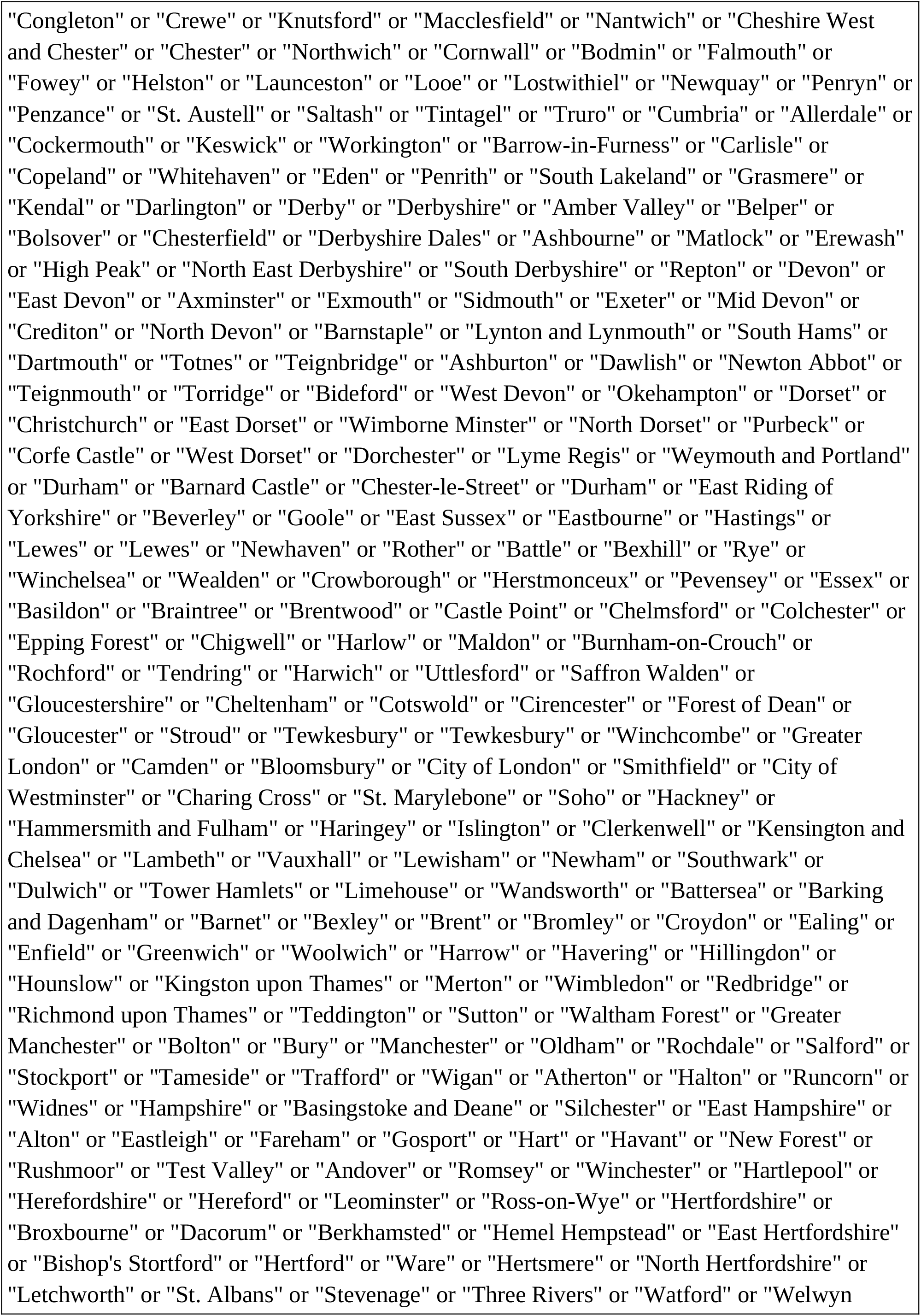
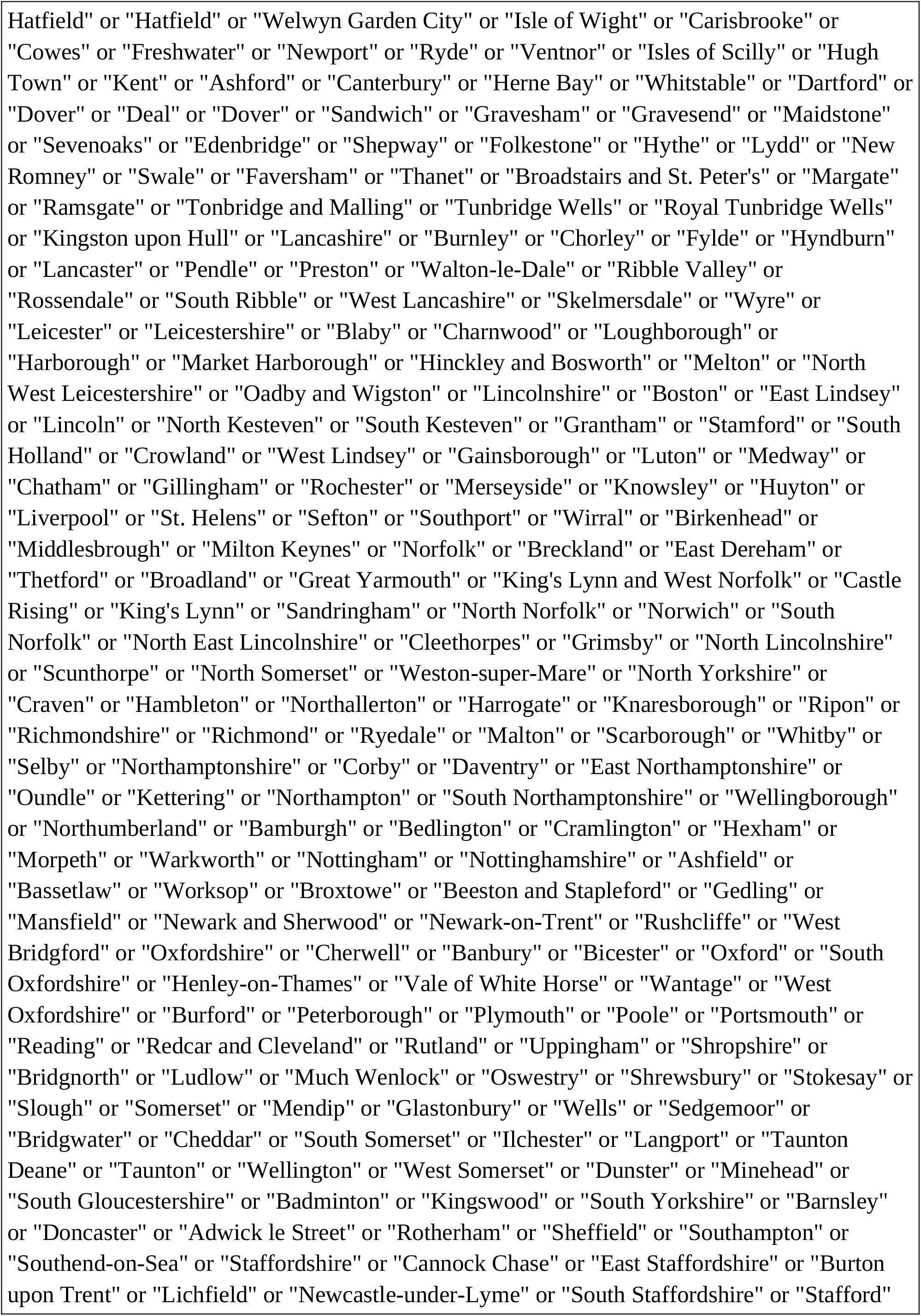
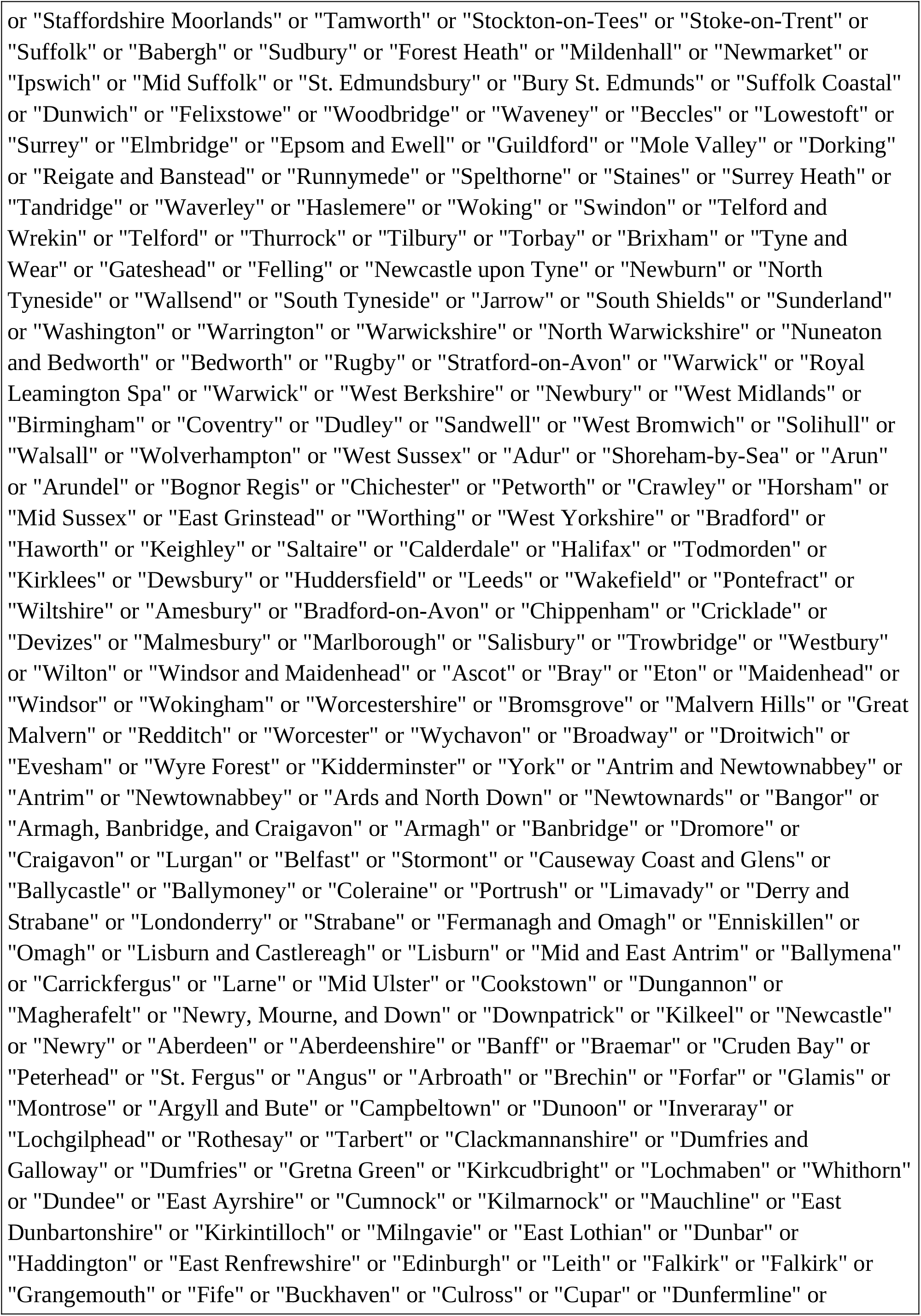
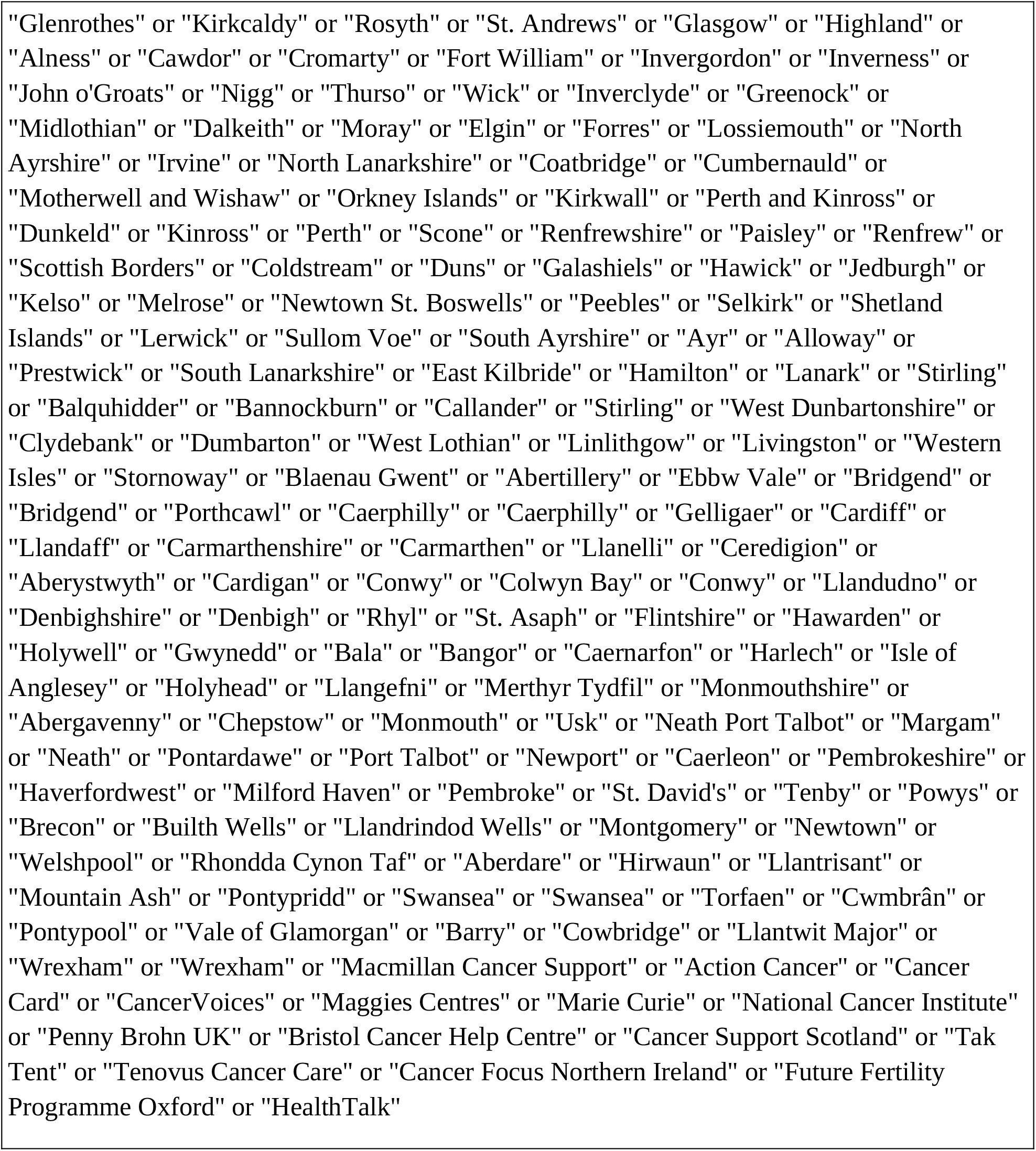
Summary of search strategy. Ovid Evidence-based Medicine Reviews (EBMR) was used to format the search, with the syntax reflecting that used in EBMR (36).

We made lists of search terms for: (**1**) the blood cancers of interest and general terms for blood cancer (*disease terms*), (**2**) the population of interest (terms for minority ethnic groups/population and ethnicity/race; note where proximity operators were not available, some of these terms were removed) (*group terms*), (**3**) haematopoietic stem cell transplant and CAR T (*therapy terms*), (**4**) UK studies (*UK terms*). See table 2 for details on search terms. The general search will be: *disease terms* AND *group terms. Therapy terms* will be used where some tools return large numbers of irrelevant references that need refining, found during piloting (EBMR Ovid, EBMR Embase, Proquest, Cochrane; added AND *therapy terms*). Where UK filters are available, these will be used (EBMR Embase, Proquest, Scopus). A custom filter for UK studies (*UK terms*), based on (37), will be used in EBMR Ovid to refine the search, where a UK filter is not available (added AND *UK terms*). Terms will be searched in the full text where possible, otherwise title/abstract. For PubMed, we will only use MESH terms separated by OR operators (MESH terms related to *disease terms* OR *group terms*).

We found some search tools lack operators that were required by our search strategy (i.e. adjacency operators; adj), which is an issue considering the complexity of the search strategy we are using. Adjacency operators look for two search terms, with a specified number of words apart. In order to ensure a sensitive search is conducted, we will programmatically (Python (40)) generate “simple searches”, from the original search, which will convert any adjacency operators to a series of phrase matching terms separated by OR. For example, where ‘(acute adj (myeloid or lymphoblastic or lymphocytic))’ is searched, the “simple search” will be ‘(“acute myeloid” OR “acute lymphoblastic” OR “acute lymphocytic”)’. This generates extremely large search strings, which will be divided into sub-searches, when using search tools that are restrictive in length of search strings.

### Study Records and Data Management

Covidence will be used to carry out and organise reviewing of the searched evidence (42). Zotero/papis will be used to organise the literature extracted after full text review (43,44). This will also be used to automate the downloading of pdfs of open-access papers. Standard Z-shell tools (45), and pdftotext (46)) will be used in the shell to convert pdfs to standard text files. These will be used as input into other tools that will be manually built to help with the screening of the papers. This will likely take the form of a simple dictionary of terms, and the count of each term in each paper (resembling simple natural language processing). Any other features that are needed, that Zotero does not provide, will be similarly handled using z-shell and Python (40,45). PRISMA flow diagram will be used to summarise the searching (18)).

### Selection Process

Two independent reviewers (SC, ZD) will conduct title and abstract screening for 500 of the abstracts, applying the exclusion/inclusion criteria outlined above. If there is a failure to reach consensus on inclusion of a study, a third party (JSC or NA) will evaluate and decide upon the verdict. If a high rate of agreement is found (>=80%), the primary author of the review (SC) will screen two thirds of the papers, and the secondary reviewer (ZD) the other third. For full text screening, both reviewers will screen 20% of the texts, and if a high rate of agreement is found (>=80%), the primary author of the review (SC) will continue independently. If the rate of agreement does not exceed this threshold, the inclusion/exclusion criteria will be clarified, and the above process repeated. If low agreement persists, then all shortlisted papers will go through full text review.

### Data Collection Process

A modified Cochrane Public Health Group Data Extraction and Assessment Template will be used for extracting data from relevant studies (47). Where any information from the studies, required by the data extraction form, is not available, the corresponding authors will be contacted to find this information. Where information remains missing, the reviewers (and third party if required), will decide whether the study is applicable to incorporate into the meta-analysis.

Similar to the full text screening, two reviewers (SC, ZD) will extract data from 20% of appropriate studies, and if a high rate of agreement is found (>=80%), the primary author of the review (SC) will continue independently. If the rate of agreement does not exceed this threshold, the data extraction form will be discussed (where differences were occurring), and the above process repeated. If low agreement persists, both reviewers will extract all appropriate data, with a third-party resolving difference in opinion (JSC, NA).

### Data Items

The data extraction tool is organised into six sections: study information, study eligibility, summary of assessment for inclusion, study details, intervention group (repeated for each group), outcomes. The outcome form consists of:

- ‘Study-level Outcome Identifier (e.g. #)’
- ‘Is there an analytic framework applied (e.g. logic model, conceptual framework)?’
- ‘Outcome definition’
- ‘Time points measured’
- ‘Time points reported’
- ‘Is there adequate latency for the outcome to be observed?’
- ‘Is the measure repeated on the same individuals or redrawn from the population/community for each time point?’
- ‘Unit of measurement (if relevant)’
- ‘Is there adequate power; uncertainty/significance measure? For scales – upper and lower limits and indicate whether high or low score is good’
- ‘How is the measure applied? Telephone survey, mail survey, in person by trained assessor, routinely collected data, other’
- ‘How is the outcome reported? Self or study assessor’
- ‘Is this outcome/tool validated?’
- ‘…And has it been used as validated?’
- ‘Is it a reliable outcome measure?’

### Risk of Bias in Individual Studies

Risk of bias will be assessed using the JBI critical appraisal tools, in which each study design (described above), has its own tool (21). This tool was used due to its wide range of study designs covered and being well-known (48). Using the same set of tools aims to retain consistency in assessing each study-type. Studies will be allocated one of three groups, as defined by the Cochrane handbook, low risk of bias, some concerns, and high risk of bias (49). Allocation to one of these groups will be decided upon by each reviewer (SC, ZD), using the results against each domain of the corresponding tool. If a consensus between reviewers is not met, further discussions between reviewers will be conducted to reach an agreement, with the use of a third party (JSC, NA), if an agreement is not met.

### Data Synthesis

Available data from included studies will be grouped into experimental and observational studies (randomised controlled trials, cohort, case-control and cross-sectional) and non-experimental/observational studies. Only results from the experimental/observational studies will be extracted for meta-analysis, whereas other studies will only be used for narrative synthesis. Only studies using the same study designs will be considered for pooling.

Due to the breadth of the study question, a wide variety of studies are expected. This means many studies may not be suitable to be pooled together in a meta-analysis. For these studies, summary tables of findings will be reported, alongside narrative synthesis. This narrative will begin with an explanation of the approach to synthesis, providing rationale for decisions made to effectively answer the research question, and assumptions made.

For the experimental/observational studies, findings will be grouped by outcome of interest (explained above). Within each group, data obtained from the group will be summarised and reported, with an emphasis on differences between outcomes of ethnic minorities compared to other ethnic groups. Data reporting outcomes by ethnicity alone will be reported first, followed by any intersectional or multi-variable results. Studies of similar UK populations will be grouped together. Ethnicity data can be defined at different levels, depending on the study objectives.

Where ethnic group results are pooled across studies, the differences between the pooled ethnic categorisations will be explicitly stated, with justifications for pooling. Decisions will be discussed between the first reviewer and the wider research team (ZD, JSC, NA, KN, AD) to reduce personal bias.

Where there are differences in study designs, a description will follow, explaining what effects this has to the interpretation of the results. Any common themes in bias from the risk of bias assessment will be explained, along with other weaknesses in study design (e.g. individuals included, representation of ethnic groups, way outcome was measured/collected) that may impact the ability to find disparities across the population of interest. Finally, the breadth of information available to address disparities in each outcome will be discussed.

For other study designs, reported data will only be used to complement the experimental/observational. This will be used to suggest reasons for outcome disparities at differing levels and provide insight into the second and third objectives of the review.

Where studies are deemed appropriate for pooling, meta-analyses will be conducted on available data, using a random-effects model, with the Hartung-Knapp-Sidik-Jonkman method (as recommended by the Cochrane handbook (50)). The level of between study variation, calculated with this method, will provide evidence on amount of information missing, which could be leading to differences in outcomes. Additionally, this method will ensure within-study differences (e.g. across study groups) will be considered. The outcomes ‘survival rates’, ‘co-morbidity risk’ (Cox hazard ratios), ‘disease burden’ and ‘comorbidity burdens’ (mean estimates), will be considered for meta-analysis. Results from meta-analyses will be visualised using forest plots. Any meta-analysis is planned to be undertaken using R (51,52) and/or Python (40).

### Meta-bias(es)

Sensitivity analyses will be conducted, looking at the effect of grey literature on any results from meta-analyses. Additionally, the effect of studies allocated as ‘some concerns’ and ‘low risk of bias’, from the risk of bias assessment, will be assessed in their effect on the results of meta-analyses. The effect of the inclusion of ‘some concerns’ will be assessed with the rest of the data, followed by ‘some concerns’ and ‘low risk of bias’. Additionally, the effect of inclusion of any data excluded due to missing information that could not be obtained from study authors (explained in data collection process above), will be assessed.

Funnel plots will be used to analyse the effect of small studies on pooled results, in addition to evidence of reporting bias. Egger test and visual inspection of plots will be used to test for asymmetry when number of studies are >=10, and studies are not similar in size (53).

Heterogeneity across pooled studies will be reported using I-squared-statistics. Tau-squared statistic will also be reported to quantify this variation, but only when number of studies are >=10 and lack of evidence of funnel plot asymmetry has been confirmed (50). This will also provide evidence to the applicability of a random-effects model over a fixed-effects model for the meta-analysis.

### Confidence in Cumulative Evidence

The strength of the identified evidence will be scored using the MINORS quality assessment tool (54).

## Data Availability

All data produced in the present study are available upon reasonable request to the authors

## Ethics and Dissemination

Once systematic review is completed, we aim to publish results in an appropriate journal (specific journal undecided). Being part of the NIHR Blood and Transplant Research Unit in Precision Transplant and Cellular Therapeutics (BTRU), outputs will be communicated with the wider group. The BTRU also has a patient partners group, in which outputs will also be communicated too.

## Author Contributions

SC, NA, JSC, KN, AD, DB conceived the project. DB, PF provided clinical expertise, especially when defining the population to be studied. SC wrote the protocol, with guidance and review from NA, JSC. SC, ZD trialled and refined the inclusion/exclusion criteria, and study aims.

## Funding Statement

This work was supported by the National Institute for Health and Care Research (NIHR) Blood and Transplant Research Unit (BTRU) in Precision Transplant and Cellular Therapeutics, University of Birmingham, UK.

## Competing Interests Statement

The authors declare that they have no known competing financial interests or personal relationships that could have appeared to influence the work reported in this paper.

## Notes

### Competing Interest Statement

The authors have declared no competing interest.

